# Spread and sources of information and misinformation about COVID-19 early during the pandemic in a U.S. national cohort study

**DOI:** 10.1101/2021.03.10.21252851

**Authors:** Drew A. Westmoreland, Amanda Berry, Rebecca Zimba, Sarah Kulkarni, Angela Parcesepe, Andrew R Maroko, Emily Poehlein, William You, Chloe Mirzayi, Shivani Kochhar, McKaylee Robertson, Levi Waldron, Christian Grov, Denis Nash

## Abstract

**Background:** Early in the pandemic, misinformation about COVID-19 was spread on social media. The purpose of this study was to describe trusted sources of COVID-19 information and claims seen and believed about COVID-19 early in the pandemic among U.S. adults. Then, we assessed the impact of believing such claims on engaging in personal protective actions (PPA).

**Methods:** We used baseline data from the CHASING COVID Cohort (*n* = 7,070) collected March 28, 2020 to April 20, 2020 to describe trusted sources of COVID-19 information as well as claims circulating on social media that had been seen and believed. We used Poisson regression to determine the association of believing certain claims with engaging in a higher number of PPA.

**Results:** The top three trusted sources of COVID-19 information were the CDC (67.9%), the WHO (53.7%), and State Health Departments (53.0%). Several COVID-19 claims circulated on social media had been seen, e.g., that the virus was created in a laboratory (54.8%). Moreover, substantial proportions of participants indicated agreement with some of these claims. In multivariable regression, we found that belief in certain claims was associated with engaging in a higher number of PPA. For example, believing that paper masks would prevent transmission of the virus was associated with engaging in a higher number of protective actions (β = 0.02, 95% CI: 0.004 – 0.046).

**Conclusions:** Results suggest the need for public health leadership on social media platforms to combat misinformation and supports social media as a tool to further public health interventions.

## INTRODUCTION

In the early days of the U.S. COVID-19 pandemic (i.e., Spring 2020), public health officials and other government entities sprang into action to reduce the impact and spread of the virus (1). While public health and medical professionals battled the virus, a new challenge emerged via misinformation spread about COVID-19 on social media. In the past decade, social media has been used proactively by public health practitioners to disseminate evidence-based public health interventions. For example, social media platforms have been used to implement HIV-testing interventions (2) and interventions aimed at reducing prescription drug misuse (3). Despite the utility of social media for disseminating evidence-based public health messaging, in recent years it has become increasingly common for social media to be co-opted by internet ne’er-do-wells with increasingly nefarious motives (4). Unfortunately, the COVID-19 pandemic did not escape this disturbing trend.

> *“But we’re not just fighting an epidemic; we’re fighting an infodemic. Fake news spreads faster and more easily than this virus, and is just as dangerous*.*”*
>
> - Dr Tedros Adhanom Ghebreyesus, Director-General, World Health Organization (5)

Nicknamed the COVID-19 “infodemic,” researchers have investigated the extent to which misinformation has circulated on social media and the patterns of content of the false messages (6, 7). In examining the content of misinformation available on COVID-19, Baines et al (2020) called the COVID-19 infodemic “unprecedented in its size and velocity” (6). Further, Broniatowski et al (2020) attempted to assess the quality of the information spread related to COVID-19, finding that most of the information could not be assessed because the amount of information originating on social media and other sources was too difficult to verify (8). Many claims that could and still can be found circulating the internet focus on rumor, stigma, and conspiracy theory (9) leading to information often being co-opted for political purposes (8) and not for the good of public health.

To our knowledge, no studies have investigated the reach of COVID-19 claims among populations participating in a public health study. Further, there is little or no data on the impact that claims on social media may have on engaging in protective public health actions during the pandemic. Using data from a U.S. national cohort during the early months of the pandemic, the aims of this paper were to: 1) describe trusted sources of COVID-19 information among cohort participants; 2) assess the reach of certain COVID-19 claims in the cohort; 3) determine the extent to which cohort participants endorsed these claims, and 4) examine the association of claims endorsements with engaging in personal protective actions.

## METHODS

Study participants were individuals screened for enrollment into the Communities, Households, and SARS-CoV-2 Epidemiology (CHASING) COVID Cohort study who completed an initial baseline assessment. The CHASING COVID Cohort study is a U.S. national prospective cohort study of adults 18 years or older that was launched on March 28, 2020 and enrolled participants through July 16, 2020 to understand the spread and impact of the SARS-CoV-2 pandemic within households and communities. The cohort methodology is described in detail elsewhere (10). Briefly, study participants included in this analysis were recruited online through social media platforms or through referrals using advertisements that were in both English and Spanish. Data was collected using Qualtrics (Qualtrics, Provo, UT), an online survey platform widely used in social and behavioral research.

This analysis uses data from the screening survey, which captured information on household characteristics, underlying risk factors, SARS-CoV-2 symptoms, and health-seeking behaviors such as testing and hospitalizations. A total of 7,070 participants completed the initial cohort screening survey between March 28, 2020 and April 20, 2020. Only one participant was recruited via Twitter and subsequently excluded from the final sample because of a very brief and failed advertising attempt on the social media site leaving 7,069 participants included in the present analyses. The study protocol was approved by the Institutional Review Board at the City University of New York (CUNY).

### Measures

From the screening survey, measures of interest to the current study were: trusted sources of COVID-19 information, exposure to certain claims about COVID-19 on social media, believing certain claims about COVID-19, actions being taken to prevent COVID-19 infection, and other covariates to help better describe the sample.

#### Sources of COVID-19 information

To determine participants’ trusted sources of COVID-19-related information, we asked participants, “[w]ho do you trust to give you reliable information regarding the new coronavirus?” Participants were able to select all that applied to them from the following list: the U.S. Centers for Disease Control and Prevention (CDC), World Health Organization (WHO), Surgeon General, White House, President, State Health Department, local (county or city) Health Department, Governor, Mayor, personal physician, other healthcare provider or worker, family member, close friend, religious leader/clergy, an online symptom checker (e.g. WebMD), significant other/spouse, work colleagues, news media, social media, other (write-in allowed), or participants could indicate that they did not trust anyone to provide reliable information regarding COVID-19.

#### Exposure to certain claims about COVID on social media

Participants were then asked, “[h]ave you seen any posts (e.g., ads, articles, or infographics) on social media (e.g., Facebook, Instagram, YouTube, Reddit) making any of the following claims about the new coronavirus?” The claims included: that the virus could be spread via a cloud of virus in the air; that the virus was created as a biological weapon; that there was a cure for the virus; and that most people who got the virus would die, in addition to other claims. The list of COVID-19-related claims in our survey was generated by reviewing claims being widely circulated on social media during March 2020 that were false, could not be substantiated, or did not come from an authoritative source, such as the CDC. Participants were asked if they had seen, had not seen, did not know/were unsure if they had seen these claims. Participants could also indicate if they were not on social media or found the questions not applicable.

#### Believing certain claims about COVID on social media

The same list of claims (as above) was then presented to participants but this time they were asked, “[t]o what extent do you agree or disagree with the following claims on social media about the coronavirus?” Participants were able to select that they strongly agreed, agreed, disagreed, or strongly disagreed with the presented claims (regardless of having seen the claim on social media). For regression analyses, a dichotomous variable was created by combining agreed and strongly agreed (herein called “agreed”) as well as disagree and strongly disagreed (herein called “disagreed”).

#### Actions taken to prevent COVID-19 infection

Participants were asked a series of questions to determine actions they had taken since the pandemic began as a result of COVID-19. The list of actions included both actions related solely to personal choice and others influenced by external forces, such as work. For the current study, we focused on those actions that involved individual decision-making only and were endorsed by public health experts as important for stemming the spread of the virus at the time of data collection. These actions were: 1) avoiding large gatherings of more than 20 people; 2) wearing a face mask; 3) increasing the frequency or length of duration of handwashing; and 4) increasing the frequency of using hand sanitizer. For each of these actions, participants were able to answer yes, no, or not applicable. These questions were then summed to estimate the total number of personal protective actions in which participants had engaged (Figure 1).

**Figure 1.**
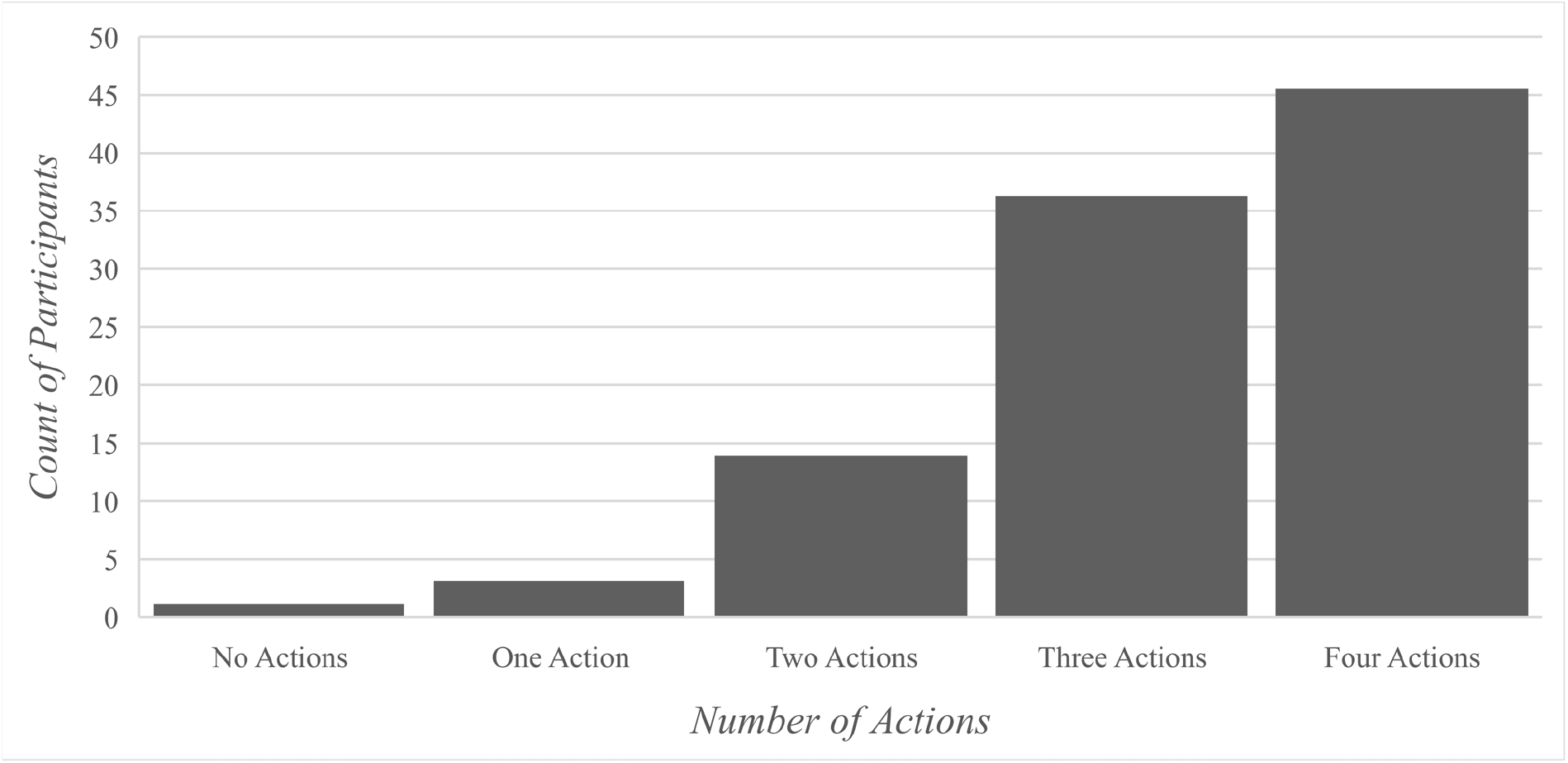
The number of personal protective actions in which participants had engaged, CHASING COVID Cohort (C3), n = 7069, 2020.

#### Covariates

There were several covariates of interest to the current study. First, this study was conducted using internet-based recruitment strategies. As such, we were interested in how the recruitment platform by which participants were recruited may have impacted where people got their trusted COVID-19 information, and exposure to and belief of certain claims. Participants were categorized based on recruitment source: Facebook, Instagram, Scruff (a social application for gay, bi, trans and queer men), marketing companies (used for targeted recruitment of certain demographics online and via various social media platforms), personal referral or networks, and through our Institute’s website. For demographic statistics, each recruitment source is presented while, for multivariable models, a dichotomous variable (n= 4,853) was created for having been recruited from social media platforms (i.e. from Facebook, Instagram, or Scruff) or not (i.e. all other recruitment sources). Respondents (n=2,154) recruited via one marketing company were excluded from the dichotomous social media platform variable, because it could not be determined where each participant saw the recruitment ad. Additionally, we include age, gender, race/ethnicity, employment status, annual income, and highest level of education as covariates.

### Analyses

Descriptive statistics (e.g., frequencies, percentages) were used to describe variables of interest. Bivariate analyses (e.g., chi-squared tests) were used to indicate differences across social media/recruitment platforms for variables of interest. Additional analyses were used to determine the number (counts) of personal protective actions in which participants had engaged. Poisson regression analyses (bivariate and multivariable) were used to assess the associations between believing certain individual claims about COVID-19 (those in which >25% of participants agreed, 7 total models) and engaging in more personal protective actions. Respondents for which it could not be determined if they were recruited via social media were excluded from these analyses (n=2,154). All multivariable models adjusted for age, gender, race/ethnicity, education status, and whether they were recruited from social media or not. For these analyses, regression coefficients, 95% confidence intervals, and p-values (α < 0.05) are reported. All analyses were conducted using SAS 9.4.

## RESULTS

The highest proportion of participants were recruited from marketing companies’ online advertising (30.8%) followed by Scruff (27.6%), our Institute’s website (17.7%), Facebook (13.3%), Instagram (7.2%), and referral/personal networks (3.0%). Fifty-one percent (51.2%) of participants identified as male and 66.6% as White, non-Hispanic. Different age groups were well represented with just over one-fifth (20.7%) being 18-29, 22.7% 30-39, 17.7% 40-49, 15.7% 50-59, and 23.2% 60 or older. Over half (52.5%) of the sample reported being employed and half (50.2%) reported making less than $50,000 a year at the time. Half (50.4%) reported having a college education. For all demographic variables, statistically significant differences were noted across recruitment platform (Table 1).

**Table 1.**
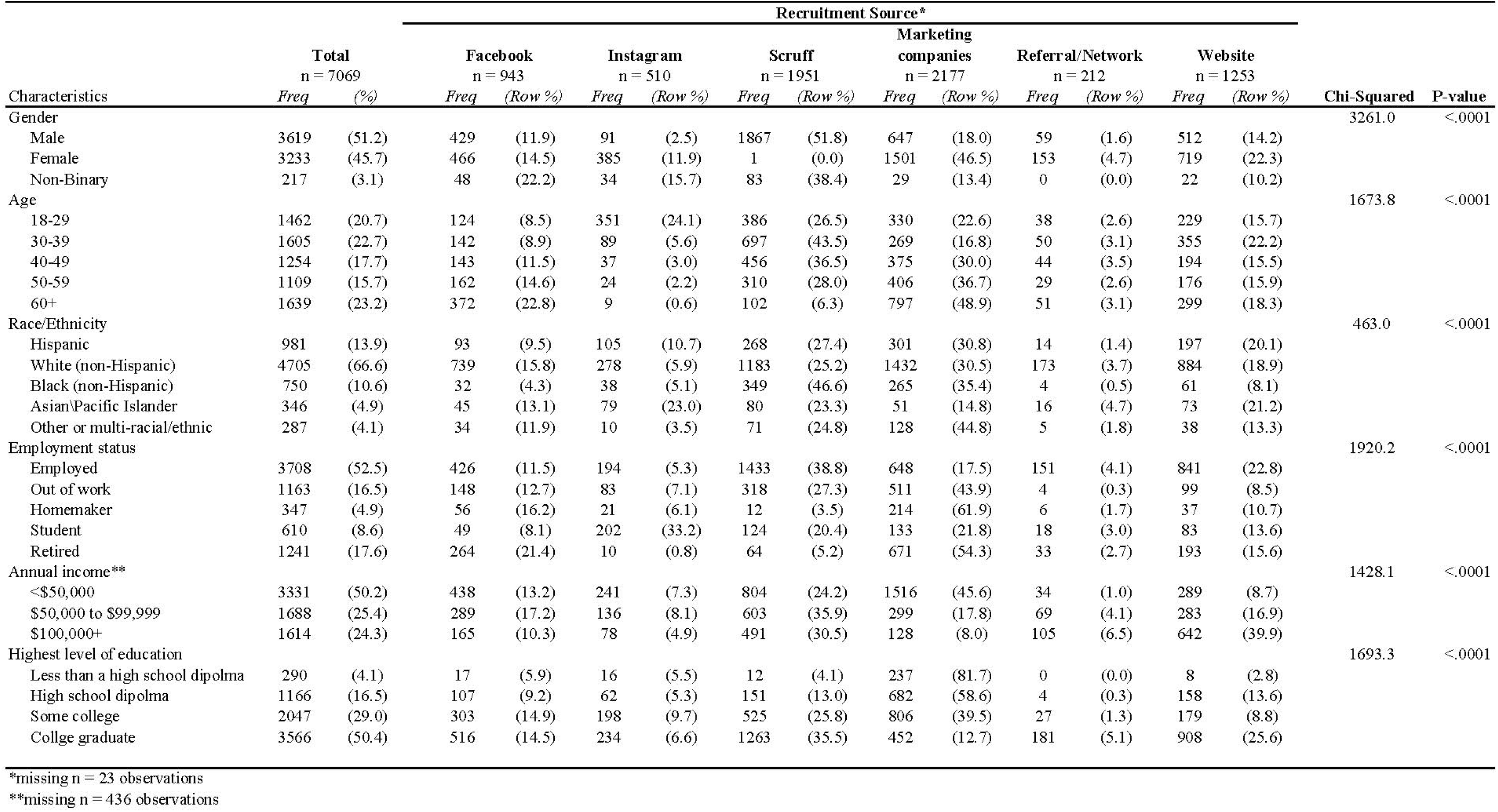
Demographic characteristics of cohort participants completing baseline and month 1 survey by recruitment source, CHASING COVID Cohort. (C3), n = 7069, 2020.

| Recruitment Source* |  |  |  |  |  |  |  |  |  |  |  |  |  |  | Chi-Squared | P-value |
| --- | --- | --- | --- | --- | --- | --- | --- | --- | --- | --- | --- | --- | --- | --- | --- | --- |
| Characteristics | Total<br>n = 7069 |  | Facebook<br>n = 943 |  | Instagram<br>n = 510 |  | Scruff<br>n = 1951 |  | Marketing<br>companies<br>n = 2177 |  | Referral/Network<br>n = 212 |  | Website<br>n = 1253 |  |  |  |
|  | Freq | (%) | Freq | (Row %) | Freq | (Row %) | Freq | (Row %) | Freq | (Row %) | Freq | (Row %) | Freq | (Row %) |  |  |
| Gender |  |  |  |  |  |  |  |  |  |  |  |  |  |  | 3261.0 | <.0001 |
| Male | 3619 | (51.2) | 429 | (11.9) | 91 | (2.5) | 1867 | (51.8) | 647 | (18.0) | 59 | (1.6) | 512 | (14.2) |  |  |
| Female | 3233 | (45.7) | 466 | (14.5) | 385 | (11.9) | 1 | (0.0) | 1501 | (46.5) | 153 | (4.7) | 719 | (22.3) |  |  |
| Non-Binary | 217 | (3.1) | 48 | (22.2) | 34 | (15.7) | 83 | (38.4) | 29 | (13.4) | 0 | (0.0) | 22 | (10.2) |  |  |
| Age |  |  |  |  |  |  |  |  |  |  |  |  |  |  | 1673.8 | <.0001 |
| 18-29 | 1462 | (20.7) | 124 | (8.5) | 351 | (24.1) | 386 | (26.5) | 330 | (22.6) | 38 | (2.6) | 229 | (15.7) |  |  |
| 30-39 | 1605 | (22.7) | 142 | (8.9) | 89 | (5.6) | 697 | (43.5) | 269 | (16.8) | 50 | (3.1) | 355 | (22.2) |  |  |
| 40-49 | 1254 | (17.7) | 143 | (11.5) | 37 | (3.0) | 456 | (36.5) | 375 | (30.0) | 44 | (3.5) | 194 | (15.5) |  |  |
| 50-59 | 1109 | (15.7) | 162 | (14.6) | 24 | (2.2) | 310 | (28.0) | 406 | (36.7) | 29 | (2.6) | 176 | (15.9) |  |  |
| 60+ | 1639 | (23.2) | 372 | (22.8) | 9 | (0.6) | 102 | (6.3) | 797 | (48.9) | 51 | (3.1) | 299 | (18.3) |  |  |
| Race/Ethnicity |  |  |  |  |  |  |  |  |  |  |  |  |  |  | 463.0 | <.0001 |
| Hispanic | 981 | (13.9) | 93 | (9.5) | 105 | (10.7) | 268 | (27.4) | 301 | (30.8) | 14 | (1.4) | 197 | (20.1) |  |  |
| White (non-Hispanic) | 4705 | (66.6) | 739 | (15.8) | 278 | (5.9) | 1183 | (25.2) | 1432 | (30.5) | 173 | (3.7) | 884 | (18.9) |  |  |
| Black (non-Hispanic) | 750 | (10.6) | 32 | (4.3) | 38 | (5.1) | 349 | (46.6) | 265 | (35.4) | 4 | (0.5) | 61 | (8.1) |  |  |
| Asian/Pacific Islander | 346 | (4.9) | 45 | (13.1) | 79 | (23.0) | 80 | (23.3) | 51 | (14.8) | 16 | (4.7) | 73 | (21.2) |  |  |
| Other or multi-racial/ethnic | 287 | (4.1) | 34 | (11.9) | 10 | (3.5) | 71 | (24.8) | 128 | (44.8) | 5 | (1.8) | 38 | (13.3) |  |  |
| Employment status |  |  |  |  |  |  |  |  |  |  |  |  |  |  | 1920.2 | <.0001 |
| Employed | 3708 | (52.5) | 426 | (11.5) | 194 | (5.3) | 1433 | (38.8) | 648 | (17.5) | 151 | (4.1) | 841 | (22.8) |  |  |
| Out of work | 1163 | (16.5) | 148 | (12.7) | 83 | (7.1) | 318 | (27.3) | 511 | (43.9) | 4 | (0.3) | 99 | (8.5) |  |  |
| Homemaker | 347 | (4.9) | 56 | (16.2) | 21 | (6.1) | 12 | (3.5) | 214 | (61.9) | 6 | (1.7) | 37 | (10.7) |  |  |
| Student | 610 | (8.6) | 49 | (8.1) | 202 | (33.2) | 124 | (20.4) | 133 | (21.8) | 18 | (3.0) | 83 | (13.6) |  |  |
| Retired | 1241 | (17.6) | 264 | (21.4) | 10 | (0.8) | 64 | (5.2) | 671 | (54.3) | 33 | (2.7) | 193 | (15.6) |  |  |
| Annual income** |  |  |  |  |  |  |  |  |  |  |  |  |  |  | 1428.1 | <.0001 |
| <\$50,000 | 3331 | (50.2) | 438 | (13.2) | 241 | (7.3) | 804 | (24.2) | 1516 | (45.6) | 34 | (1.0) | 289 | (8.7) | | |
| \$50,000 to \$99,999 | 1688 | (25.4) | 289 | (17.2) | 136 | (8.1) | 603 | (35.9) | 299 | (17.8) | 69 | (4.1) | 283 | (16.9) | | |
| \$100,000+ | 1614 | (24.3) | 165 | (10.3) | 78 | (4.9) | 491 | (30.5) | 128 | (8.0) | 105 | (6.5) | 642 | (39.9) | | |
| Highest level of education |  |  |  |  |  |  |  |  |  |  |  |  |  |  | 1693.3 | <.0001 |
| Less than a high school diploma | 290 | (4.1) | 17 | (5.9) | 16 | (5.5) | 12 | (4.1) | 237 | (81.7) | 0 | (0.0) | 8 | (2.8) |  |  |
| High school diploma | 1166 | (16.5) | 107 | (9.2) | 62 | (5.3) | 151 | (13.0) | 682 | (58.6) | 4 | (0.3) | 158 | (13.6) |  |  |
| Some college | 2047 | (29.0) | 303 | (14.9) | 198 | (9.7) | 525 | (25.8) | 806 | (39.5) | 27 | (1.3) | 179 | (8.8) |  |  |
| College graduate | 3566 | (50.4) | 516 | (14.5) | 234 | (6.6) | 1263 | (35.5) | 452 | (12.7) | 181 | (5.1) | 908 | (25.6) |  |  |

\*missing n = 23 observations
\*\*missing n = 436 observations

### Sources of information about COVID-19

Across the total sample, the top three sources of information about COVID-19 (Table 2) were the CDC (67.9%), the WHO (53.7%), and the participants’ State Health Departments (53.0%). Participants who were recruited on Instagram, Scruff, referral/personal networks, and the ISPH website had the same top three trusted sources. Differences were noted for participants recruited from Facebook (72.2% CDC, 58.0% WHO, 57.1% personal physician) and the marketing companies (50.8% CDC, 42.7% personal physician, 40.7% State Health Department).

**Table 2.** Trusted sources of COVID-19 information and with whom participants discussed COVID-19, CHASING COVID Cohort (C3), *n* = 7069, 2020.

|  | Recruitment Source* |  |  |  |  |  |  |  |  |  |  |  |  |  |
| --- | --- | --- | --- | --- | --- | --- | --- | --- | --- | --- | --- | --- | --- | --- |
|  | Total<br>n = 7069 |  | Facebook<br>n = 943 |  | Instagram<br>n = 510 |  | Scruff<br>n = 1951 |  | Marketing companies<br>n = 2177 |  | Referral/Network<br>n = 212 |  | Website<br>n = 1253 |  |
|  | Freq | (%) | Freq | (Col %) | Freq | (Col %) | Freq | (Col %) | Freq | (Col %) | Freq | (Col %) | Freq | (Col %) |
| Who do you trust to give you reliable information regarding the new coronavirus? |  |  |  |  |  |  |  |  |  |  |  |  |  |  |
| CDC | 4796 | (67.9) | 681 | (72.2) | 403 | (79.0) | 1512 | (77.5) | 1106 | (50.8) | 167 | (78.8) | 909 | (72.6) |
| WHO | 3798 | (53.7) | 547 | (58.0) | 377 | (73.9) | 1329 | (68.1) | 567 | (26.1) | 151 | (71.2) | 813 | (64.9) |
| State Health Department | 3747 | (53.0) | 524 | (55.6) | 309 | (60.6) | 1168 | (59.9) | 886 | (40.7) | 134 | (63.2) | 714 | (57.0) |
| Personal physician | 3599 | (50.9) | 538 | (57.1) | 214 | (42.0) | 1111 | (57.0) | 929 | (42.7) | 125 | (59.0) | 671 | (53.6) |
| Local/County/City Health Department | 3508 | (49.6) | 507 | (53.8) | 281 | (55.1) | 1080 | (55.4) | 806 | (37.0) | 123 | (58.0) | 700 | (55.9) |
| Your governor | 2805 | (39.7) | 414 | (43.9) | 191 | (37.5) | 843 | (43.2) | 613 | (28.2) | 127 | (59.9) | 607 | (48.4) |
| Other healthcare provider/worker | 2635 | (37.3) | 388 | (41.2) | 204 | (40.0) | 834 | (42.8) | 579 | (26.6) | 96 | (45.3) | 525 | (41.9) |
| Surgeon General | 2132 | (30.2) | 322 | (34.2) | 197 | (38.6) | 578 | (29.6) | 585 | (26.9) | 65 | (30.7) | 378 | (30.2) |
| News media | 2030 | (28.7) | 277 | (29.4) | 147 | (28.8) | 642 | (32.9) | 471 | (21.6) | 90 | (42.5) | 399 | (31.8) |
| Family member | 1402 | (19.8) | 172 | (18.2) | 82 | (16.1) | 293 | (15.0) | 573 | (26.3) | 43 | (20.3) | 236 | (18.8) |
| Your mayor | 1354 | (19.2) | 179 | (19.0) | 97 | (19.0) | 486 | (24.9) | 298 | (13.7) | 45 | (21.2) | 245 | (19.6) |
| Significant other/spouse | 1142 | (16.2) | 169 | (17.9) | 67 | (13.1) | 209 | (10.7) | 383 | (17.6) | 61 | (28.8) | 250 | (20.0) |
| Close friend | 1055 | (14.9) | 105 | (11.1) | 61 | (12.0) | 303 | (15.5) | 354 | (16.3) | 36 | (17.0) | 192 | (15.3) |
| President | 910 | (12.9) | 110 | (11.7) | 32 | (6.3) | 96 | (4.9) | 588 | (27.0) | 7 | (3.3) | 74 | (5.9) |
| Online symptom checker | 756 | (10.7) | 125 | (13.3) | 55 | (10.8) | 189 | (9.7) | 232 | (10.7) | 24 | (11.3) | 129 | (10.3) |
| White House | 712 | (10.1) | 87 | (9.2) | 38 | (7.5) | 110 | (5.6) | 389 | (17.9) | 6 | (2.8) | 80 | (6.4) |
| Work colleagues | 557 | (7.9) | 48 | (5.1) | 24 | (4.7) | 180 | (9.2) | 106 | (4.9) | 30 | (14.2) | 167 | (13.3) |
| Social media | 531 | (7.5) | 64 | (6.8) | 46 | (9.0) | 115 | (5.9) | 223 | (10.2) | 7 | (3.3) | 75 | (6.0) |
| Religious leader/clergy | 385 | (5.5) | 41 | (4.4) | 11 | (2.2) | 49 | (2.5) | 229 | (10.5) | 2 | (0.9) | 52 | (4.2) |
| None of the above | 327 | (4.6) | 48 | (5.1) | 17 | (3.3) | 71 | (3.6) | 163 | (7.5) | 0 | (0.0) | 28 | (2.2) |
| Who do you talk to about the new coronavirus? |  |  |  |  |  |  |  |  |  |  |  |  |  |  |
| Immediate family | 5452 | (77.1) | 741 | (78.6) | 451 | (88.4) | 1603 | (82.2) | 1449 | (66.6) | 195 | (92.0) | 996 | (79.5) |
| Close friend, someone you see or talk to regularly | 4933 | (69.8) | 610 | (64.7) | 389 | (76.3) | 1631 | (83.6) | 1148 | (52.7) | 190 | (89.6) | 951 | (75.9) |
| Partner/significant other | 3965 | (56.1) | 615 | (65.2) | 289 | (56.7) | 798 | (40.9) | 1191 | (54.7) | 169 | (79.7) | 887 | (70.8) |
| Extended/other family | 2848 | (40.3) | 333 | (35.3) | 163 | (32.0) | 792 | (40.6) | 724 | (33.3) | 122 | (57.6) | 703 | (56.1) |
| Coworker | 2572 | (36.4) | 257 | (27.3) | 151 | (29.6) | 1118 | (57.3) | 337 | (15.5) | 124 | (58.5) | 580 | (46.3) |
| Acquaintance, someone you know, but don't see or talk to often | 2055 | (29.1) | 239 | (25.3) | 159 | (31.2) | 841 | (43.1) | 361 | (16.6) | 79 | (37.3) | 371 | (29.6) |
| Medical provider or other health care professional | 2055 | (29.1) | 268 | (28.4) | 92 | (18.0) | 609 | (31.2) | 670 | (30.8) | 66 | (31.1) | 344 | (27.5) |
| I don't feel comfortable talking about it with anyone | 212 | (3.0) | 19 | (2.0) | 11 | (2.2) | 41 | (2.1) | 130 | (6.0) | 0 | (0.0) | 11 | (0.9) |

\*missing *n* = 23 observations

When examining who people talked to about COVID-19 (Table 2), the top three responses were immediate family (77.1%), close friend/someone you see or talk to regularly (69.8%), and partner/significant other (56.1%). Participants recruited from Facebook, Instagram, the marketing companies, referral/personal networks, and the ISPH website also had similar patterns of with whom they discussed COVID-19. The only difference was for participants recruited on Scruff; their top three for discussing COVID-19 included their co-workers (57.3%), and did not include partner/significant other.

### COVID-19 claims seen by participants

In total, over one-third (37.2%) of participants had seen the claim that the virus could be spread via “a cloud in the air” (Supplemental Table 1). Over half (54.8%) had seen the claim that the virus was created in a laboratory and that the virus was created as part of a biological weapon (51.6%). Just over a third (35.1%) of participants had seen the claim that home remedies and cures would cure the virus, but fewer (15.2%) had seen the claim that ingesting or bathing in disinfectants would cure the virus. Slightly over one-fifth (21.2%) had seen claims that there was a cure for the virus. Forty-three (43.1%) percent had seen that disposable paper masks would prevent getting the virus and over half (55.9%) of participants had seen that heat could kill the virus. Over one-third (35.3%) had seen claims that children could not get the virus, 14.3% had seen claims that all or most people who got the virus would die, 28.1% had seen claims that people who “recovered” from the virus could suddenly become ill again and die, and 39.8% had seen that pets can get the virus. Lastly, over one-fifth (22.8%) of participants had seen claims that ordering products from China could make them sick.

Across all recruitment platforms, there were differences in the proportion of participants who had seen certain claims. A few key results are presented in-text. Full results are presented in Supplemental Table 1. Three-quarters (75.3%) of participants recruited from Facebook reported that they had seen the claim that the virus was created in a laboratory and 70.0% reported that they had seen claims the virus was created as part of a biological weapon. In general, for these two claims, higher proportions of participants reported that they had seen the claims on social media (compared to those who had not did not know/were unsure if they had seen claims, or were not on social media) across all recruitment methods with the two exceptions being for personal referrals (for both claims). Conversely, across most recruitment platforms, a majority of participants reported that they had not seen claims that there was a cure for the virus.

### Participants who believed COVID-19 claims

Regardless of whether participants had seen the above claims on social media, they were asked their level of agreement with all claims. Nearly half (47.2%) of participants agreed with the claim that the virus could be spread via “a cloud in the air” (Supplemental Table 1). About one-third (32.6%) agreed that the virus had been created in a laboratory and just over one-quarter (26.4%) agreed with claims that the virus had been created as a part of a biological weapon. Fewer participants indicated that they agreed with claims that home remedies (11.8%) or ingesting/bathing in disinfectants (7.6%) could cure the virus. Just over one-fifth (21.5%) of participants reported that they agreed with the claim that there was a cure for the virus. Twenty-nine (29.4%) percent agreed with the claim that disposable paper masks could prevent getting the virus, and 43.5% of participants agreed with the claim that heat can kill the virus. Eleven (11.1%) percent agreed with claims that children could not get the virus, 9.7% agreed with claims that all or most people who got the virus would die, 29.3% agreed with claims that people who “recovered” from the virus could suddenly become ill again and die, and 34.7% agreed with claims that pets can get the virus. Finally, 12.8% of participants agreed with claims that ordering products from China would make them sick.

As with exposure to claims on social media, there was variability in the proportions of participants who agreed with the claims between recruitment methods. All results are presented in Supplemental Table 1, but a few results are highlighted here. Across the total sample and most recruitment platforms, lower proportions of participants agreed with the claim that ordering products from China would make them sick. However, 25.9% of participants recruited via marketing companies endorsed this claim compared to only 11.1% of participants recruited through Facebook or 6.3% from Scruff, for example. Even for claims endorsed by more of the total sample, higher proportions of participants recruited via marketing companies indicated agreement. For example, 59.2% of participants recruited using marketing companies agreed with claims that the virus was created in a laboratory (32.7% of total sample) and 49.9% agreed/strongly agreed with claims that the virus was created as part of a biological weapon (26.4% of total sample). Comparatively, 18.1% and 17.3% of participants recruited through Instagram agreed with these claims, respectively.

#### Association of claims beliefs with engaging in protective actions

Finally, we used simple bivariate and multivariable Poisson regression analyses (7 total models, n=4,853) to determine if believing in certain claims impacted the total number of individual protective actions in which participants engaged (Table 3). Multivariable results suggest that agreeing with claims that the virus can be spread via “a cloud of virus in the air” (β = 0.04, 95% CI 0.004 – 0.068, *p*-value = 0.03), disposable paper masks will prevent you from getting the virus (β = 0.02, 95% CI 0.004 – 0.046, *p*-value = 0.02), many people who recover from the virus can suddenly become ill again and die (β = 0.03, 95% CI 0.005 – 0.05, *p*-value = 0.02), and pets can get the virus and become ill (β = 0.03, 95% CI 0.007 – 0.046, *p*-value = 0.01) were all associated with engaging in a higher number of personal protective actions.

**Table 3.**
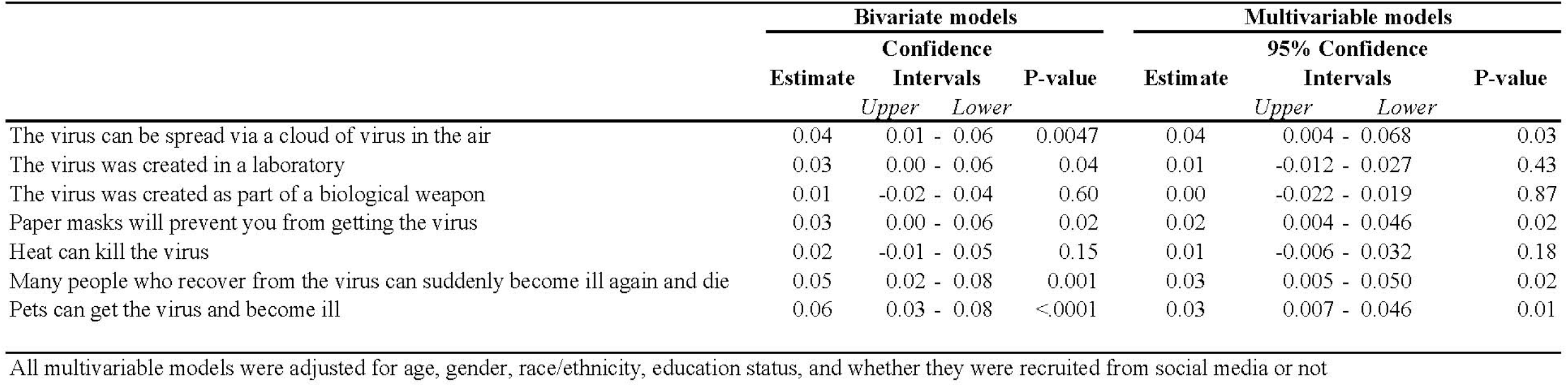
Bivariate and multivariable associations of agreeing with certain COVID-19 claims on social media with increasing number of personal protective actions, CHASING COVID Cohort (C3), n = 4853, 2020.

## DISCUSSION

The main goals of this study were to describe participants’ trusted sources of COVID-19-related information, determine which select claims about COVID-19 people had seen circulating on social media, the extent to which people agreed with these claims, and whether agreeing with these claims impacted engaging in personal protective actions. The three most common sources of trusted COVID-19-related information were the CDC, the WHO, and State Health Departments. Participants reported discussing COVID-19 with a variety of individuals but most commonly their immediate family, close friends, and partners/significant others. Several COVID-19 claims had been seen on social media including claims that the virus was created in a laboratory and that children cannot get the virus. Further, substantial proportions of participants indicated agreeing or strongly agreeing with some of these claims. Finally, we also found that agreeing with certain claims increased the number of personal protective actions in which participants engaged.

In the early months of the U.S. pandemic, participants reported that their trusted sources of information regarding COVID-19 were the CDC, the WHO, and State Health Departments. These agencies are responsible for managing public health emergencies, including pandemics, and it is encouraging that participants viewed these agencies as trusted sources for COVID-19 information. While the WHO’s reputation suffered after the West African Ebola epidemic of 2013-2016 (11), the CDC has been a long-recognized global leader in public health (12). However, early mistakes and continued blunders have led to a litany of newspaper articles criticizing the agency’s COVID-19 response (13, 14), and have compromised the CDC’s reputation as a global and national leader in public health. Future research among U.S. residents should continue to evaluate the reputations of the CDC and WHO (15)— especially given the recent politization of the U.S.’s membership in the WHO, the recent negative press coverage of the CDC, and new leadership in the White House and other federal agencies—as their reputations are important for advancing evidence-based public health policies and programs. If trust in the CDC and WHO has decreased since the beginning of the COVID-19 pandemic, State Health Departments could leverage their reputations to continue providing U.S. residents with correct and timely public health information.

Although our findings regarding trusted sources of information are encouraging, our findings regarding claims circulating on social media about COVID-19 highlight the proportions of participants encountering these claims on social media and those vulnerable to believing misinformation. Social connections and social media are useful sources of health information (16). However, studies into patterns of information sharing behavior on social media help to illuminate how complicated the motivation of social media behavior can be. Individuals can be influenced by motivations other than sharing accurate, factual information, such as a desire for social community conformity or harmony, that impact the articles, videos, and postings they choose to share (17, 18). People can also take cues from “opinion leaders” who can direct followers to certain sources of information and even change followers’ own personal views (19). These opinion leaders can be particularly influential, especially if they have elevated status such as “celebrity”, regardless of their level of expertise (20). Understanding this dynamic is especially important when considering the spread of misinformation and differing approaches to how and when opinion leaders and information seekers validate the information they are sharing or reading, where opinion leaders are more likely to validate and information seekers are less likely (19).

When looking at the infodemic impacting COVID-19 responses, public health professionals and advocates were attempting to combat misinformation grounded in conspiracy theory and political machinations (8, 21) with evolving scientific evidence. Scientific processes can sometimes undermine the authority of public health professionals (e.g., debates on e-cigarettes and vaping), and careless presentation of study findings can lead to stigmatization and misinformation (e.g., HIV/AIDS crisis) (22). Evolving and conflicting scientific evidence is a necessary part of scientific discovery. The need to make informed decisions despite evolving scientific evidence is a reality in nearly every pandemic due to a novel pathogen. The absence of stable evidence may leave individuals more susceptible to the harmful effects of misinformation. Therefore, with or without clear and complete evidence to support the public health response, public health professionals and advocates should become more familiar with the underlying motivations driving misinformation on social media so that we can become better public health “opinion leaders” as well as experts.

Finally, our results provide some evidence that believing certain claims on social media was associated with engaging in personal protective actions. Multivariable regression analyses indicated that certain claims circulating early during the pandemic—notably believing that COVID-19 was spread via a “cloud in the air” and that disposable masks could prevent the virus—were associated with engaging in a higher count of preventive actions. While the current research supports aerosolized transmission (23) and that masks can prevent transmission of the virus (24), early research, at the time of this survey, was not conclusive. However, both claims, at their core, address concerns over transmissibility of the virus. Our study suggests these claims related to the spread of the virus were associated with engaging in more personal protective behaviors while other claims related to the virus’ origins were not found to be associated with engaging in personal protective actions. Our findings indicating persons engaging in more personal protective actions is encouraging as an indication of how seriously some people took the pandemic early on despite a lack of consistent messaging from trusted sources on how the virus was being transmitted and best-practice protective measures. Some claims assessed in this study were “conspiratorial” in nature and some, including purported preventive measures and cures, were harmful as evidenced by news media from around the globe (9, 25). Future research needs to continue evaluating the impacts of social media misinformation on COVID-19-related health behaviors as well as other health outcomes during the pandemic.

Our study should be interpreted in light of its limitations. First, our study used multiple recruitment methods to achieve sampling goals (e.g., age and racial distributions). As such, participants were recruited from methods other than social media and resulting participant samples across available social media platforms are not equal. Our recruitment strategies limit comparisons between different social media platforms in terms of what claims were seen. However, they do help to highlight the importance of considering social media platforms when recruiting participants for research studies. Further, we did not ask on which social media platform participants had seen certain claims. This limits our ability to see which claims were circulating on certain platforms, however this data collection would be subject to recall bias as many people are on multiple platforms.

The CHASING COVID Cohort was established early in the pandemic’s spread throughout the U.S., and these data are taken from the baseline assessment. Unlike other countries, the U.S. failed to “flatten” its curve, seeing a second surge in the summer, and a third in the fall 2020 (at the time this manuscript was prepared). It is important to understand our results in the context of when the data were collected. It would also be important to understand how trustworthiness of COVID-19 information may have changed over time given shifting views of how the public views the pandemic itself (e.g., pandemic fatigue) as well as governmental response (or lack thereof) to control the pandemic’s spread.

## CONCLUSIONS

Across recruitment sources, the CDC, WHO, and State Health Departments tended to be the most trusted sources for COVID-19 information, and immediate family members, close friends, and partners tended to be the persons with whom participants talked about the pandemic. For the most part, participants did not agree with many incorrect claims about the pandemic, such as bathing or ingesting chemical disinfectants as a means of prevention/cure. Nevertheless, and often while in the minority, an alarming number of individuals did agree with inaccurate claims about the pandemic. Misinformation spread on social media and other outlets needs to be immediately addressed by trusted sources of health information by counter campaigns highlighting the needs for a strong social media and online presence. For future pandemics, it will be important to identify ways in which social media could be leveraged to help ensure that the most accurate and evidence-based information gets reinforced, and misinformation spread is reduced or prevented, in support of public health efforts.

## Data Availability

De-identified data may be made available upon request.

## ACKNOWLEDGEMENTS

We would like to acknowledge the CHASING COVID Cohort Study participants for their contributions to this research.

---

**Supplementary Table 1.** Claims seen circulating on social media about COVID-19 by social media recruitment source, CHASING COVID Cohort (C3), n = 7069, 2020.

| Recruitment Source |  |  |  |  |  |  |  |  |  |  |  |  |  |  |  |  |  |  |
| --- | --- | --- | --- | --- | --- | --- | --- | --- | --- | --- | --- | --- | --- | --- | --- | --- | --- | --- |
|  | Total<br>n = 7,069 |  | Facebook<br>n = 943 |  | Instagram<br>n = 510 |  | Scruff<br>n = 1951 |  | Twitter<br>n = 1 |  | Marketing companies<br>n = 2177 |  | Referral/Network<br>n = 212 |  | Web site<br>n = 1253 |  |  |  |
|  | Freq | (Col %) | Freq | (Col %) | Freq | (Col %) | Freq | (Col %) | Freq | (Col %) | Freq | (Col %) | Freq | (Col %) | Freq | (Col %) | Chi-squared | p |
| Seen claims |  |  |  |  |  |  |  |  |  |  |  |  |  |  |  |  |  |  |
| The virus can be spread via a cloud of virus in the air |  |  |  |  |  |  |  |  |  |  |  |  |  |  |  |  | 328.52 | < .0001 |
| Have not seen | 2918 | (41.4) | 366 | (38.9) | 231 | (45.3) | 871 | (44.7) | 0 | (0.0) | 833 | (38.7) | 102 | (48.1) | 508 | (40.5) |  |  |
| Have seen | 2619 | (37.2) | 467 | (49.6) | 215 | (42.2) | 719 | (36.9) | 0 | (0.0) | 696 | (32.3) | 75 | (35.4) | 433 | (34.6) |  |  |
| Don't know/Not sure | 846 | (12.0) | 86 | (9.1) | 53 | (10.4) | 189 | (9.7) | 1 | (100.0) | 390 | (18.1) | 13 | (6.1) | 113 | (9.0) |  |  |
| Not on social media/NA | 660 | (9.4) | 22 | (2.3) | 11 | (2.2) | 171 | (8.8) | 0 | (0.0) | 235 | (10.9) | 22 | (10.4) | 199 | (15.9) |  |  |
| The virus was created in a laboratory |  |  |  |  |  |  |  |  |  |  |  |  |  |  |  |  | 524.29 | < .0001 |
| Have not seen | 1962 | (27.9) | 158 | (16.8) | 137 | (26.9) | 662 | (34.0) | 0 | (0.0) | 525 | (24.5) | 103 | (48.6) | 373 | (29.8) |  |  |
| Have seen | 3850 | (54.8) | 709 | (75.3) | 332 | (65.1) | 1039 | (53.3) | 1 | (100.0) | 1042 | (48.7) | 85 | (40.1) | 625 | (49.9) |  |  |
| Don't know/Not sure | 757 | (10.8) | 46 | (4.9) | 25 | (4.9) | 107 | (5.5) | 0 | (0.0) | 410 | (19.1) | 3 | (1.4) | 165 | (13.2) |  |  |
| Not on social media/NA | 462 | (6.6) | 29 | (3.1) | 16 | (3.1) | 141 | (7.2) | 0 | (0.0) | 165 | (7.7) | 21 | (9.9) | 90 | (7.2) |  |  |
| The virus was created as part of a biological weapon |  |  |  |  |  |  |  |  |  |  |  |  |  |  |  |  | 535.45 | < .0001 |
| Have not seen | 2128 | (30.3) | 204 | (21.7) | 139 | (27.3) | 662 | (34.0) | 1 | (100.0) | 616 | (28.7) | 98 | (46.2) | 403 | (32.2) |  |  |
| Have seen | 3631 | (51.6) | 659 | (70.0) | 330 | (64.7) | 1044 | (53.6) | 0 | (0.0) | 895 | (41.8) | 87 | (41.0) | 600 | (47.9) |  |  |
| Don't know/Not sure | 806 | (11.5) | 57 | (6.1) | 22 | (4.3) | 103 | (5.3) | 0 | (0.0) | 445 | (20.8) | 6 | (2.8) | 172 | (13.7) |  |  |
| Not on social media/NA | 466 | (6.6) | 21 | (2.2) | 19 | (3.7) | 140 | (7.2) | 0 | (0.0) | 187 | (8.7) | 21 | (9.9) | 78 | (6.2) |  |  |
| Home remedies such as eating garlic or sesame seed oil and using essential oils |  |  |  |  |  |  |  |  |  |  |  |  |  |  |  |  |  |  |
| colloidal silver zinc and steroids will cure the virus |  |  |  |  |  |  |  |  |  |  |  |  |  |  |  |  | 674.65 | < .0001 |
| Have not seen | 3252 | (46.3) | 394 | (41.9) | 234 | (45.9) | 963 | (49.4) | 0 | (0.0) | 974 | (45.5) | 115 | (54.3) | 562 | (44.9) |  |  |
| Have seen | 2470 | (35.1) | 458 | (48.7) | 229 | (44.9) | 761 | (39.1) | 1 | (100.0) | 500 | (23.3) | 73 | (34.4) | 437 | (34.9) |  |  |
| Don't know/Not sure | 687 | (9.8) | 58 | (6.2) | 31 | (6.1) | 96 | (4.9) | 0 | (0.0) | 446 | (20.8) | 3 | (1.4) | 52 | (4.2) |  |  |
| Not on social media/NA | 620 | (8.8) | 31 | (3.3) | 16 | (3.1) | 129 | (6.6) | 0 | (0.0) | 222 | (10.4) | 21 | (9.9) | 201 | (16.1) |  |  |
| Ingesting or bathing in chemical disinfectants will prevent or cure the virus |  |  |  |  |  |  |  |  |  |  |  |  |  |  |  |  | 481.02 | < .0001 |
| Have not seen | 4703 | (66.9) | 641 | (68.1) | 371 | (72.8) | 1388 | (71.2) | 0 | (0.0) | 1273 | (59.5) | 167 | (78.8) | 847 | (67.7) |  |  |
| Have seen | 1071 | (15.2) | 215 | (22.9) | 101 | (19.8) | 338 | (17.3) | 1 | (100.0) | 233 | (10.9) | 19 | (9.0) | 160 | (12.8) |  |  |
| Don't know/Not sure | 711 | (10.1) | 49 | (5.2) | 28 | (5.5) | 76 | (3.9) | 0 | (0.0) | 393 | (18.4) | 5 | (2.4) | 158 | (12.6) |  |  |
| Not on social media/NA | 543 | (7.7) | 36 | (3.8) | 10 | (2.0) | 147 | (7.5) | 0 | (0.0) | 242 | (11.3) | 21 | (9.9) | 87 | (7.0) |  |  |
| There is a cure for the virus |  |  |  |  |  |  |  |  |  |  |  |  |  |  |  |  | 605.88 | < .0001 |
| Have not seen | 4376 | (62.3) | 583 | (62.0) | 326 | (63.9) | 1318 | (67.7) | 0 | (0.0) | 1163 | (54.3) | 164 | (77.4) | 801 | (64.0) |  |  |
| Have seen | 1488 | (21.2) | 293 | (31.1) | 149 | (29.2) | 411 | (21.1) | 1 | (100.0) | 400 | (18.7) | 24 | (11.3) | 209 | (16.7) |  |  |
| Don't know/Not sure | 600 | (8.5) | 37 | (3.9) | 24 | (4.7) | 89 | (4.6) | 0 | (0.0) | 399 | (18.6) | 1 | (0.5) | 50 | (4.0) |  |  |
| Not on social media/NA | 564 | (8.0) | 28 | (3.0) | 11 | (2.2) | 130 | (6.7) | 0 | (0.0) | 180 | (8.4) | 23 | (10.9) | 192 | (15.3) |  |  |
| Paper masks will prevent you from getting the virus |  |  |  |  |  |  |  |  |  |  |  |  |  |  |  |  | 391.83 | < .0001 |
| Have not seen | 2678 | (38.1) | 393 | (41.8) | 193 | (37.8) | 818 | (42.0) | 0 | (0.0) | 661 | (30.8) | 107 | (50.5) | 497 | (39.7) |  |  |
| Have seen | 3026 | (43.1) | 457 | (48.6) | 269 | (52.8) | 874 | (44.8) | 1 | (100.0) | 847 | (39.5) | 77 | (36.3) | 488 | (39.0) |  |  |
| Don't know/Not sure | 939 | (13.4) | 70 | (7.4) | 42 | (8.2) | 138 | (7.1) | 0 | (0.0) | 484 | (22.6) | 9 | (4.3) | 196 | (15.7) |  |  |
| Not on social media/NA | 386 | (5.5) | 21 | (2.2) | 6 | (1.2) | 119 | (6.1) | 0 | (0.0) | 151 | (7.1) | 19 | (9.0) | 70 | (5.6) |  |  |
| Heat can kill the virus |  |  |  |  |  |  |  |  |  |  |  |  |  |  |  |  | 535.30 | < .0001 |
| Have not seen | 2069 | (29.5) | 196 | (20.9) | 148 | (29.0) | 552 | (28.3) | 0 | (0.0) | 655 | (30.7) | 66 | (31.1) | 448 | (35.9) |  |  |
| Have seen | 3922 | (55.9) | 684 | (72.8) | 322 | (63.1) | 1184 | (60.8) | 1 | (100.0) | 918 | (43.0) | 120 | (56.6) | 676 | (54.1) |  |  |
| Don't know/Not sure | 651 | (9.3) | 39 | (4.2) | 29 | (5.7) | 102 | (5.2) | 0 | (0.0) | 416 | (19.5) | 5 | (2.4) | 59 | (4.7) |  |  |
| Not on social media/NA | 377 | (5.4) | 21 | (2.2) | 11 | (2.2) | 110 | (5.7) | 0 | (0.0) | 148 | (6.9) | 21 | (9.9) | 66 | (5.3) |  |  |
| Kids can't get the virus |  |  |  |  |  |  |  |  |  |  |  |  |  |  |  |  | 483.83 | < .0001 |
| Have not seen | 3485 | (49.6) | 462 | (49.2) | 256 | (50.2) | 1085 | (55.7) | 1 | (100.0) | 992 | (46.4) | 109 | (51.4) | 573 | (45.8) |  |  |
| Have seen | 2478 | (35.3) | 409 | (43.6) | 225 | (44.1) | 657 | (33.7) | 0 | (0.0) | 635 | (29.7) | 80 | (37.7) | 457 | (36.5) |  |  |
| Don't know/Not sure | 523 | (7.5) | 48 | (5.1) | 20 | (3.9) | 82 | (4.2) | 0 | (0.0) | 333 | (15.6) | 2 | (0.9) | 38 | (3.0) |  |  |
| Not on social media/NA | 535 | (7.6) | 20 | (2.1) | 9 | (1.8) | 124 | (6.4) | 0 | (0.0) | 178 | (8.3) | 21 | (9.9) | 183 | (14.6) |  |  |
| All or most people who get the virus will die |  |  |  |  |  |  |  |  |  |  |  |  |  |  |  |  | 740.89 | < .0001 |
| Have not seen | 4853 | (69.1) | 751 | (79.9) | 381 | (74.7) | 1528 | (78.4) | 0 | (0.0) | 1068 | (50.0) | 177 | (83.5) | 928 | (74.2) |  |  |
| Have seen | 1003 | (14.3) | 120 | (12.8) | 93 | (18.2) | 224 | (11.5) | 0 | (0.0) | 456 | (21.3) | 12 | (5.7) | 95 | (7.6) |  |  |
| Don't know/Not sure | 768 | (10.9) | 46 | (4.9) | 30 | (5.9) | 85 | (4.4) | 1 | (100.0) | 448 | (21.0) | 1 | (0.5) | 158 | (12.6) |  |  |
| Not on social media/NA | 397 | (5.7) | 23 | (2.5) | 6 | (1.2) | 111 | (5.7) | 0 | (0.0) | 166 | (7.8) | 22 | (10.4) | 69 | (5.5) |  |  |
| Many people who recover from the virus can suddenly become ill again and die |  |  |  |  |  |  |  |  |  |  |  |  |  |  |  |  | 426.17 | < .0001 |
| Have not seen | 3943 | (56.2) | 490 | (52.1) | 310 | (60.8) | 1222 | (62.7) | 0 | (0.0) | 1024 | (48.0) | 134 | (63.2) | 752 | (60.1) |  |  |
| Have seen | 1970 | (28.1) | 363 | (38.6) | 157 | (30.8) | 501 | (25.7) | 1 | (100.0) | 531 | (24.9) | 51 | (24.1) | 356 | (28.5) |  |  |
| Don't know/Not sure | 767 | (10.9) | 66 | (7.0) | 34 | (6.7) | 123 | (6.3) | 0 | (0.0) | 460 | (21.6) | 8 | (3.8) | 75 | (6.0) |  |  |
| Not on social media/NA | 338 | (4.8) | 21 | (2.2) | 9 | (1.8) | 102 | (5.2) | 0 | (0.0) | 119 | (5.6) | 19 | (9.0) | 68 | (5.4) |  |  |
| Ordering products from China will make me sick |  |  |  |  |  |  |  |  |  |  |  |  |  |  |  |  | 639.32 | < .0001 |
| Have not seen | 4158 | (59.3) | 538 | (57.2) | 319 | (62.6) | 1290 | (66.3) | 1 | (100.0) | 1031 | (48.4) | 158 | (74.5) | 806 | (64.5) |  |  |
| Have seen | 1598 | (22.8) | 326 | (34.6) | 154 | (30.2) | 456 | (23.4) | 0 | (0.0) | 416 | (19.5) | 27 | (12.7) | 212 | (17.0) |  |  |
| Don't know/Not sure | 851 | (12.1) | 56 | (6.0) | 27 | (5.3) | 88 | (4.5) | 0 | (0.0) | 513 | (24.1) | 8 | (3.8) | 159 | (12.7) |  |  |
| Not on social media/NA | 408 | (5.8) | 21 | (2.2) | 10 | (2.0) | 113 | (5.8) | 0 | (0.0) | 172 | (8.1) | 19 | (9.0) | 73 | (5.8) |  |  |
| Pets can get the virus and become ill |  |  |  |  |  |  |  |  |  |  |  |  |  |  |  |  | 732.77 | < .0001 |
| Have not seen | 3166 | (45.1) | 396 | (42.1) | 219 | (42.9) | 1097 | (56.4) | 0 | (0.0) | 742 | (34.8) | 122 | (57.6) | 580 | (46.4) |  |  |
| Have seen | 2790 | (39.8) | 483 | (51.3) | 263 | (51.6) | 677 | (34.8) | 1 | (100.0) | 849 | (39.8) | 70 | (33.0) | 436 | (34.9) |  |  |
| Don't know/Not sure | 592 | (8.4) | 41 | (4.4) | 22 | (4.3) | 73 | (3.8) | 0 | (0.0) | 406 | (19.0) | 1 | (0.5) | 49 | (3.9) |  |  |
| Not on social media/NA | 467 | (6.7) | 21 | (2.2) | 6 | (1.2) | 99 | (5.1) | 0 | (0.0) | 135 | (6.3) | 19 | (9.0) | 186 | (14.9) |  |  |
| *Believe claims* |  |  |  |  |  |  |  |  |  |  |  |  |  |  |  |  |  |  |
| The virus can be spread via a cloud of virus in the air |  |  |  |  |  |  |  |  |  |  |  |  |  |  |  |  | 252.77 | < .0001 |
| Strongly Disagree | 1039 | (14.8) | 123 | (13.1) | 99 | (19.5) | 377 | (19.4) | 0 | (0.0) | 198 | (9.2) | 33 | (15.7) | 204 | (16.4) |  |  |
| Disagree | 2672 | (38.1) | 330 | (35.2) | 217 | (42.7) | 740 | (38.1) | 0 | (0.0) | 726 | (33.8) | 97 | (46.2) | 556 | (44.6) |  |  |
| Agree | 2369 | (33.8) | 358 | (38.2) | 154 | (30.3) | 583 | (30.0) | 1 | (100.0) | 818 | (38.1) | 71 | (33.8) | 377 | (30.3) |  |  |
| Strongly Agree | 937 | (13.4) | 126 | (13.5) | 38 | (7.5) | 245 | (12.6) | 0 | (0.0) | 406 | (18.9) | 9 | (4.3) | 109 | (8.8) |  |  |
| The virus was created in a laboratory |  |  |  |  |  |  |  |  |  |  |  |  |  |  |  |  | 1392.56 | < .0001 |
| Strongly Disagree | 2232 | (31.9) | 328 | (34.9) | 221 | (43.4) | 806 | (41.4) | 0 | (0.0) | 185 | (8.7) | 120 | (57.1) | 561 | (45.1) |  |  |
| Disagree | 2487 | (35.5) | 362 | (38.6) | 196 | (38.5) | 735 | (37.8) | 0 | (0.0) | 685 | (32.1) | 80 | (38.1) | 424 | (34.1) |  |  |
| Agree | 1556 | (22.2) | 161 | (17.2) | 63 | (12.4) | 292 | (15.0) | 1 | (100.0) | 811 | (38.0) | 7 | (3.3) | 217 | (17.4) |  |  |
| Strongly Agree | 731 | (10.4) | 88 | (9.4) | 29 | (5.7) | 113 | (5.8) | 0 | (0.0) | 453 | (21.2) | 3 | (1.4) | 43 | (3.5) |  |  |
| The virus was created as part of a biological weapon |  |  |  |  |  |  |  |  |  |  |  |  |  |  |  |  | 1372.53 | < .0001 |
| Strongly Disagree | 2439 | (34.8) | 376 | (40.0) | 237 | (46.6) | 850 | (43.7) | 0 | (0.0) | 218 | (10.2) | 131 | (62.4) | 615 | (49.4) |  |  |
| Disagree | 2714 | (38.8) | 366 | (39.0) | 184 | (36.2) | 720 | (37.0) | 1 | (100.0) | 850 | (39.9) | 69 | (32.9) | 519 | (41.7) |  |  |
| Agree | 1226 | (17.5) | 135 | (14.4) | 64 | (12.6) | 264 | (13.6) | 0 | (0.0) | 677 | (31.8) | 9 | (4.3) | 73 | (5.9) |  |  |
| Strongly Agree | 624 | (8.9) | 62 | (6.6) | 24 | (4.7) | 112 | (5.8) | 0 | (0.0) | 385 | (18.1) | 1 | (0.5) | 39 | (3.1) |  |  |
| Home remedies such as eating garlic or sesame seed oil and using essential oils colloidal silver zinc and steroids will cure the virus |  |  |  |  |  |  |  |  |  |  |  |  |  |  |  |  | 848.25 | < .0001 |
| Strongly Disagree | 3283 | (46.9) | 504 | (53.5) | 285 | (56.0) | 1055 | (54.3) | 0 | (0.0) | 490 | (23.0) | 131 | (62.7) | 802 | (64.2) |  |  |
| Disagree | 2892 | (41.3) | 354 | (37.6) | 179 | (35.2) | 695 | (35.8) | 1 | (100.0) | 1189 | (55.9) | 73 | (34.9) | 396 | (31.7) |  |  |
| Agree | 610 | (8.7) | 60 | (6.4) | 35 | (6.9) | 147 | (7.6) | 0 | (0.0) | 323 | (15.2) | 5 | (2.4) | 39 | (3.1) |  |  |
| Strongly Agree | 217 | (3.1) | 24 | (2.6) | 10 | (2.0) | 46 | (2.4) | 0 | (0.0) | 125 | (5.9) | 0 | (0.0) | 12 | (1.0) |  |  |
| Ingesting or bathing in chemical disinfectants will prevent or cure the virus |  |  |  |  |  |  |  |  |  |  |  |  |  |  |  |  | 1001.15 | < .0001 |
| Strongly Disagree | 4190 | (59.9) | 651 | (69.3) | 370 | (72.7) | 1411 | (72.5) | 1 | (100.0) | 731 | (34.4) | 160 | (76.6) | 846 | (67.7) |  |  |
| Disagree | 2283 | (32.6) | 252 | (26.8) | 127 | (25.0) | 482 | (24.8) | 0 | (0.0) | 1102 | (51.9) | 47 | (22.5) | 272 | (21.8) |  |  |
| Agree | 368 | (5.3) | 21 | (2.2) | 9 | (1.8) | 30 | (1.5) | 0 | (0.0) | 181 | (8.5) | 2 | (1.0) | 124 | (9.9) |  |  |
| Strongly Agree | 160 | (2.3) | 16 | (1.7) | 3 | (0.6) | 22 | (1.1) | 0 | (0.0) | 111 | (5.2) | 0 | (0.0) | 8 | (0.6) |  |  |
| There is a cure for the virus |  |  |  |  |  |  |  |  |  |  |  |  |  |  |  |  | 802.45 | < .0001 |
| Strongly Disagree | 2375 | (33.9) | 371 | (39.5) | 166 | (32.6) | 827 | (42.6) | 0 | (0.0) | 329 | (15.5) | 115 | (54.8) | 552 | (44.2) |  |  |
| Disagree | 3120 | (44.6) | 408 | (43.4) | 254 | (49.9) | 778 | (40.1) | 1 | (100.0) | 1006 | (47.3) | 90 | (42.9) | 578 | (46.3) |  |  |
| Agree | 1139 | (16.3) | 119 | (12.7) | 80 | (15.7) | 251 | (12.9) | 0 | (0.0) | 591 | (27.8) | 5 | (2.4) | 91 | (7.3) |  |  |
| Strongly Agree | 366 | (5.2) | 42 | (4.5) | 9 | (1.8) | 85 | (4.4) | 0 | (0.0) | 202 | (9.5) | 0 | (0.0) | 28 | (2.2) |  |  |
| Paper masks will prevent you from getting the virus |  |  |  |  |  |  |  |  |  |  |  |  |  |  |  |  | 278.73 | < .0001 |
| Strongly Disagree | 1447 | (20.7) | 243 | (25.9) | 116 | (22.8) | 500 | (25.8) | 0 | (0.0) | 279 | (13.1) | 52 | (24.8) | 249 | (20.0) |  |  |
| Disagree | 3492 | (49.9) | 480 | (51.1) | 253 | (49.7) | 918 | (47.3) | 1 | (100.0) | 1006 | (47.3) | 112 | (53.3) | 711 | (57.0) |  |  |
| Agree | 1760 | (25.2) | 195 | (20.7) | 132 | (25.9) | 453 | (23.3) | 0 | (0.0) | 672 | (31.6) | 44 | (21.0) | 261 | (20.9) |  |  |
| Strongly Agree | 297 | (4.2) | 22 | (2.3) | 8 | (1.6) | 70 | (3.6) | 0 | (0.0) | 169 | (8.0) | 2 | (1.0) | 26 | (2.1) |  |  |
| Heat can kill the virus |  |  |  |  |  |  |  |  |  |  |  |  |  |  |  |  | 115.88 | < .0001 |
| Strongly Disagree | 1063 | (15.2) | 143 | (15.2) | 93 | (18.3) | 353 | (18.2) | 0 | (0.0) | 239 | (11.3) | 40 | (19.1) | 188 | (15.1) |  |  |
| Disagree | 2884 | (41.3) | 384 | (40.9) | 221 | (43.4) | 742 | (38.3) | 1 | (100.0) | 945 | (44.7) | 97 | (46.4) | 490 | (39.4) |  |  |
| Agree | 2450 | (35.1) | 337 | (35.9) | 172 | (33.8) | 691 | (35.6) | 0 | (0.0) | 685 | (32.4) | 63 | (30.1) | 493 | (39.6) |  |  |
| Strongly Agree | 583 | (8.4) | 76 | (8.1) | 23 | (4.5) | 153 | (7.9) | 0 | (0.0) | 246 | (11.6) | 9 | (4.3) | 74 | (5.9) |  |  |
| Kids can't get the virus |  |  |  |  |  |  |  |  |  |  |  |  |  |  |  |  | 603.48 | < .0001 |
| Strongly Disagree | 3414 | (48.8) | 493 | (52.5) | 290 | (57.0) | 1164 | (60.0) | 1 | (100.0) | 653 | (30.8) | 124 | (59.1) | 673 | (53.8) |  |  |
| Disagree | 2799 | (40.0) | 374 | (39.8) | 188 | (36.9) | 651 | (33.5) | 0 | (0.0) | 989 | (46.7) | 78 | (37.1) | 515 | (41.2) |  |  |
| Agree | 470 | (6.7) | 40 | (4.3) | 23 | (4.5) | 78 | (4.0) | 0 | (0.0) | 287 | (13.5) | 5 | (2.4) | 36 | (2.9) |  |  |
| Strongly Agree | 309 | (4.4) | 33 | (3.5) | 8 | (1.6) | 48 | (2.5) | 0 | (0.0) | 190 | (9.0) | 3 | (1.4) | 26 | (2.1) |  |  |
| All or most people who get the virus will die |  |  |  |  |  |  |  |  |  |  |  |  |  |  |  |  | 866.41 | < .0001 |
| Strongly Disagree | 3114 | (44.6) | 514 | (54.7) | 233 | (45.8) | 1059 | (54.6) | 1 | (100.0) | 464 | (21.9) | 144 | (68.6) | 682 | (54.6) |  |  |
| Disagree | 3196 | (45.7) | 376 | (40.0) | 237 | (46.6) | 771 | (39.7) | 0 | (0.0) | 1221 | (57.7) | 63 | (30.0) | 525 | (42.0) |  |  |
| Agree | 449 | (6.4) | 30 | (3.2) | 32 | (6.3) | 80 | (4.1) | 0 | (0.0) | 279 | (13.2) | 2 | (1.0) | 24 | (1.9) |  |  |
| Strongly Agree | 231 | (3.3) | 20 | (2.1) | 7 | (1.4) | 31 | (1.6) | 0 | (0.0) | 154 | (7.3) | 1 | (0.5) | 18 | (1.4) |  |  |
| Many people who recover from the virus can suddenly become ill again and die |  |  |  |  |  |  |  |  |  |  |  |  |  |  |  |  | 311.07 | < .0001 |
| Strongly Disagree | 1182 | (16.9) | 138 | (14.7) | 111 | (21.8) | 459 | (23.7) | 0 | (0.0) | 213 | (10.1) | 51 | (24.4) | 208 | (16.7) |  |  |
| Disagree | 3750 | (53.7) | 506 | (53.8) | 264 | (51.9) | 1076 | (55.5) | 1 | (100.0) | 1089 | (51.5) | 123 | (58.9) | 682 | (54.7) |  |  |
| Agree | 1790 | (25.6) | 263 | (28.0) | 119 | (23.4) | 365 | (18.8) | 0 | (0.0) | 661 | (31.3) | 34 | (16.3) | 338 | (27.1) |  |  |
| Strongly Agree | 259 | (3.7) | 34 | (3.6) | 15 | (3.0) | 39 | (2.0) | 0 | (0.0) | 150 | (7.1) | 1 | (0.5) | 19 | (1.5) |  |  |
| Ordering products from China will make me sick |  |  |  |  |  |  |  |  |  |  |  |  |  |  |  |  | 1035.82 | < .0001 |
| Strongly Disagree | 2891 | (41.4) | 440 | (46.8) | 261 | (51.3) | 1081 | (55.7) | 1 | (100.0) | 357 | (16.9) | 130 | (62.2) | 606 | (48.5) |  |  |
| Disagree | 3204 | (45.9) | 397 | (42.2) | 210 | (41.3) | 739 | (38.1) | 0 | (0.0) | 1207 | (57.2) | 74 | (35.4) | 572 | (45.8) |  |  |
| Agree | 661 | (9.5) | 83 | (8.8) | 31 | (6.1) | 93 | (4.8) | 0 | (0.0) | 398 | (18.8) | 5 | (2.4) | 49 | (3.9) |  |  |
| Strongly Agree | 229 | (3.3) | 21 | (2.2) | 7 | (1.4) | 29 | (1.5) | 0 | (0.0) | 150 | (7.1) | 0 | (0.0) | 22 | (1.8) |  |  |
| Pets can get the virus and become ill |  |  |  |  |  |  |  |  |  |  |  |  |  |  |  |  | 667.11 | < .0001 |
| Strongly Disagree | 1662 | (23.8) | 256 | (27.2) | 121 | (23.8) | 668 | (34.4) | 0 | (0.0) | 210 | (10.0) | 74 | (35.2) | 326 | (26.2) |  |  |
| Disagree | 2896 | (41.5) | 413 | (43.9) | 213 | (41.9) | 816 | (42.1) | 1 | (100.0) | 795 | (37.7) | 96 | (45.7) | 554 | (44.5) |  |  |
| Agree | 2000 | (28.7) | 225 | (23.9) | 153 | (30.1) | 383 | (19.7) | 0 | (0.0) | 857 | (40.6) | 38 | (18.1) | 338 | (27.1) |  |  |
| Strongly Agree | 421 | (6.0) | 47 | (5.0) | 22 | (4.3) | 73 | (3.8) | 0 | (0.0) | 248 | (11.8) | 2 | (1.0) | 28 | (2.3) |  |  |

\*missing n = 23 observations

## Notes

### Competing Interest Statement

The authors have declared no competing interest.

### Clinical Trial

This is not an intervention study.

### Clinical Protocols

https://www.medrxiv.org/content/10.1101/2020.04.28.20080630v2#p-5

### Funding Statement

Funding for this project is provided by the CUNY Institute for Implementation Science in Population Health (cunyisph.org), the COVID-19 Grant Program of the CUNY Graduate School of Public Health and Health Policy, and the National Institute Of Allergy and Infectious Diseases of the National Institutes of Health under Award Number UH3AI133675. The NIH played no role in the production of this manuscript nor necessarily endorses the findings.

### Author Declarations

The study protocol was approved by the Institutional Review Board at the City University of New York (CUNY) Graduate School for Public Health and Health Policy.

